# The impact of the COVID-19 pandemic on Antidepressant Prescribing with a focus on people with learning disability and autism: An interrupted time-series analysis in England using OpenSAFELY-TPP

**DOI:** 10.1101/2024.05.08.24306990

**Authors:** Christine Cunningham, Orla Macdonald, Andrea L Schaffer, Andrew Brown, Milan Wiedemann, Rose Higgins, Chris Bates, John Parry, Louis Fisher, Helen J Curtis, Amir Mehrkar, Liam C Hart, Seb Bacon, William Hulme, Victoria Speed, Thomas Ward, Richard Croker, Chris Wood, Alex Walker, Colm Andrews, Ben Butler-Cole, Dave Evans, Peter Inglesby, Iain Dillingham, Simon Davy, Lucy Bridges, Tom O’Dwyer, Steve Maude, Rebecca Smith, Ben Goldacre, Brian MacKenna

## Abstract

**Background:** COVID-19 lockdowns led to increased reports of depressive symptoms in the general population and impacted the health and social care services of people with learning disability and autism. We explored whether the COVID-19 pandemic had an impact on antidepressant prescribing trends within these and the general population.

**Methods:** With the approval of NHS England, we used >24 million patients’ primary care data from the OpenSAFELY-TPP platform. We identified patients with learning disability or autism and used an interrupted time series analysis to quantify trends in those prescribed and newly prescribed an antidepressant across key demographic and clinical subgroups, comparing pre-COVID-19 (January 2018-February 2020), COVID-19 lockdown (March 2020-February 2021) and the recovery period (March 2021-December 2022).

**Results:** Prior to COVID-19 lockdown, antidepressant prescribing was increasing at 0.3% (95% CI 0.2% to 0.3%) patients per month, in the general population and in those with learning disability, and 0.3% (95% CI 0.2% to 0.4%) in those with autism. We did not find evidence that the pandemic was associated with a change in trend of antidepressant prescribing in the general population (RR 1.00 (95% CI 0.97 to 1.02)), in those with autism (RR 0.99 (95% CI 0.97 to 1.01)), or in those with learning disability (RR 0.98 (95% CI 0.96 to 1.00)).

New prescribing post lockdown was 13% and 12% below expected if COVID-19 had not happened in both the general population and those with autism (RR 0.87 (95% CI 0.83 to 0.93), RR 0.88 (95% CI 0.83 to 0.92))), but not learning disability (RR 0.96 (95% CI 0.87 to 1.05)).

**Conclusions and Implications:** Pre-COVID-19, antidepressant prescribing was increasing at 0.3% per month. While we did not see an impact of COVID-19 on overall prescribing in the general population, prescriptions to those aged 0-19, 20-29, and new prescriptions were lower than pre-COVID-19 trends would have predicted, but tricyclics and new prescriptions in care homes were higher than expected.

**What is already known on this topic:**

⇒ The prescribing of antidepressants in the UK has been increasing for more than a decade.
⇒ Studies globally have found differing impacts of COVID-19 on mental health outcomes in the general population, by age, sex, socio-economic status, and care home status.

**What this study adds:**

⇒ This study describes the impact of COVID-19 on antidepressant prescribing in England with additional follow-up through December 2022, with a focus on people with a learning disability or autism.

**How this study might affect research, practice, or policy:**

⇒ This study demonstrates how the pandemic did not lead to an increase in antidepressant prescriptions in the general population, but more is needed to ensure that antidepressants are used appropriately within vulnerable populations.
⇒ Improvements are needed in the documentation of diagnosis when prescribing medicines.

## Background

The prescribing of antidepressants in the UK has been increasing for more than a decade [1,2]. During the COVID-19 pandemic, the Office of National Statistics Opinions and Lifestyle Survey found that the proportion of individuals reporting depressive symptoms during lockdown was close to double pre-pandemic levels [3]. Yet, concerns have been raised about potential overprescribing of antidepressants, particularly to those with mild depression [4,5].

In 2016, the Stopping Over Medication of People with a learning disability, autism or both (STOMP) initiative was introduced to reduce inappropriate or overprescribing in those groups [6]. Starting in the 2019-2020 year, NHS Digital (NHSD) introduced indicators to support STOMP, including tracking the proportion of patients with learning disability prescribed an antidepressant without an active depression diagnosis. Comparing 2020-2021 to 2016-2017, they found an 0.8% increase from 10.8% to 11.6% in this metric [7]. During the COVID-19 pandemic, some services offered to people with learning disability or autism and their families and carers were suspended or restricted [8], and literature from that period has described the negative impact that the lockdowns had on people with learning disability and autism, such as increases in anxiety and depressive symptoms [9–11].

OpenSAFELY is a secure analytics platform for electronic patient records built by our group on behalf of NHS England to deliver urgent academic [12–14] and operational research during the pandemic [15–17]. It allows researchers to rapidly assess the effect of the pandemic on health related outcomes. All code and analysis is shared openly for inspection and re-use.

We set out to use the OpenSAFELY platform to assess the impact of the COVID-19 pandemic on antidepressant prescribing in patients in the general population, those with learning disability and/or autism and in key demographic and clinical subgroups.

## Methods

### Study design

With the approval of NHS England, we performed an interrupted time series analysis (ITSA) to examine changes in monthly rates of the number of registered patients prescribed an antidepressant for the five year period January 2018-December 2022. We compared the 26 months from January 2018 to February 2020 (pre COVID-19), 12 months from March 2020 to February 2021 (lockdown), and 22 months from March 2021 to December 2022 (recovery).

The UK government announced its COVID response plan on March 3rd, 2020, advised against non-essential travel on March 16th, and legal restrictions came into force on March 26th [18,19], with subsequent lockdowns starting from October 31, 2020 and January 4, 2021. Exposure to the lockdown period was defined as starting in March 2020 through February 2021. The recovery period was defined as starting in March of 2021 as primary and secondary schools reopened on March 8, 2021, suggesting a return to some essential activities. This also allowed for the lockdown period to include one full calendar year. The ITSA allowed us to use the respective populations as their own control, taking into account pre-existing trends (the counterfactual) and seasonality in antidepressant prescribing, comparing these against the actual prescribing trends that occurred during and after the pandemic [20].

### Data Source and processing

The dataset analysed within OpenSAFELY is based on > 24 million people currently registered with GP surgeries using TPP SystmOne software. All data were linked, stored and analysed securely using the OpenSAFELY platform, https://www.opensafely.org/, as part of the NHS England OpenSAFELY COVID-19 service. Data include pseudonymised data such as coded diagnoses, medications and physiological parameters. No free text data are included. All code is shared openly for review and re-use under MIT open licence. Detailed pseudonymised patient data is potentially re-identifiable and therefore not shared. Data management and analysis was performed using Python 3. Code for data management and analysis, as well as codelists, are archived online at https://github.com/opensafely/antidepressant-prescribing-lda. As EQUATOR guidelines for ITSA (CARITS) are still under development, the Jandoc et al recommendations were followed [21].

### Study population

We included all individuals who were alive and registered at an OpenSAFELY-TPP practice each month, across the study period. Those with unknown age and sex were excluded as their small numbers would have necessitated redactions of the next smallest group to avoid potential re-identification of individuals, and their relative percentages are described in **Table S1**.

### Study outcomes

Antidepressants were classified as medicines falling under British National Formulary (BNF) codes starting with 040303 (selective serotonin reuptake inhibitor (SSRI)), 040301 (tricyclic), 040302 (monoamine-oxidase inhibitors (MAOI)), and other 040304 (other antidepressant). For the tricyclic and other codelists, some specific codes were excluded as these medications are more commonly used for indications other than depression. Full codelists and details on the codelist development methodology can be found on www.opencodelists.org. Links to codelists for primary outcomes used in the final study can be found in **Table S2**.

In this study, counts represent the number of patients with at least one antidepressant prescription issued in that time period. Patients with more than one antidepressant prescription in a month were only counted once. This analysis reflects prescribing rather than dispensing or use.

### Population Characteristics

We characterised all patients prescribed an antidepressant in October 2022, as a representative month close to the end of the study period. Prescribing in November and December has been shown to be subject to more seasonal variation [22]. Antidepressant prescribing rates were expressed as the number of patients prescribed an antidepressant per 1,000 registered patients. Patient counts of <=5 were redacted with remaining counts rounded to the nearest 10 to avoid potential re-identification of patients.

In October 2022 we also counted the number of patients (and relative percentage) prescribed each antidepressant type. MAOI and other antidepressants were combined into a single “other” group. Those prescribed two different classes of antidepressants in the single month (e.g. tricyclic and MAOI or SSRI and other) are counted separately as “multiple.”

Learning disability and autism diagnoses were defined using the NHSD primary care reference set codelists. These codelists are developed by NHSD for use in the Quality and Outcomes Framework (QOF) business rules, which contain indicators of good clinical care and general practitioners (GPs) can receive financial incentives based on their achievement of certain thresholds [23]. The learning disability and autism groups were non-exclusive (e.g., patients with both learning disability and autism were counted in both groups). From 2020-2021, NHSD found 28.6% of those with learning disability also had a diagnosis of autism [24].

Additional variables were defined for age (0-19, 20-29, 30-39, 40-49, 50-59, 60-69, 70-79, and 80+ years), sex, 2019 Index of Multiple Deprivation (IMD) as quintiles, ethnicity according to the 2001 census (White, Mixed, Asian or Asian British, Black or Black British, Chinese or Other, or Unknown), the 9 English geographic regions (North East, North West, Yorkshire and The Humber, East Midlands, West Midlands, East, London, South East, and South West) and care home status. The OpenSAFELY-TPP population has been shown to be broadly representative of the English population, but there are some regional coverage differences due to Electronic Health Record (EHR) system use, with the highest coverage in the East of England (91%) and lower coverage in London (19%) [25]. Care should be taken when interpreting regional rates, as they could reflect other demographic differences.

Diagnosis groups were defined as whether a patient was on the QOF depression register (active depression diagnosis), had an anxiety diagnosis, both, or neither. QOF business rules for the depression register include logic to exclude resolved diagnoses [23]. The anxiety codelist was developed based on a keyword search for “anxiety” or “anxious” within listed SNOMED CT codes. Resolved codes for anxiety were not considered, and a diagnosis could have occurred at any time in the past. Inclusion/exclusion details can be found on opencodelists (**Table S1**). The diagnosis measure is limited to those aged 18 and older, as per the QOF register definition. Children were not included for this part of the analysis.

Patient demographics were analysed monthly, except ethnicity, which for reduced computational time used the latest recorded code for each patient, as it does not often change over time [26].

### Antidepressant prescribing during the COVID-19 pandemic

Our primary aim was to understand whether the interruptions caused by the COVID-19 lockdown impacted antidepressant prescribing in England, and the impact on the at-risk learning disability and autism populations. Secondary aims were to assess the impact in other demographic or clinical subgroups.

#### Overall prescribing

Counts of patients with any antidepressant prescription per month were modelled with a Poisson regression. The log of the total population was included as an offset term to compute a rate rather than a count, given that the registered population was increasing over time.

The model included variables representing the pre-COVID-19 trend (slope), a step and slope change for the COVID-19 lockdown, and a step and slope change for the start of the recovery period. March 2020 and April 2020 appeared to be extreme outliers and were coded as dummy variables. One pair of Fourier terms were included to adjust for seasonality.

To account for autocorrelation and heteroscedasticity, robust standard errors were computed using the Newey West method [27]. Newey West corrected errors also allowed use of a Poisson model rather than a Negative Binomial model as Poisson estimates are valid in the presence of robust standard errors [28], even if there is overdispersion.

ITSA were illustrated with scatter plots of the observed data, a line showing the model fitted data, and a dotted line showing the no COVID-19 counterfactual. The counterfactual was computed by using the fitted model to estimate the rate with the step, slope and outlier variables set to 0.

An overall estimate of whether post-lockdown prescribing was below the no COVID-19 counterfactual was computed based on a method proposed by Travis-Lumer et al. [29,30]. A relative risk (RR) comparing the model fitted value to the no COVID-19 counterfactual was computed for each time point using the variance covariance matrix, and then the geometric mean of those values was computed to produce an overall effect size for the entire post intervention time period. To give an estimate of an absolute effect, we separately report the difference between the predicted and counterfactual rates and confidence intervals in the last month of the study period.

#### New prescribing

We also analysed the number of patients “newly” prescribed an antidepressant. This model similarly counted the number of patients prescribed an antidepressant each month, but the denominator was those registered patients without any antidepressant prescription for the past 2 years, or “antidepressant naive.”

#### Learning Disability and Autism subgroups

For each month, we identified those with a current or previous diagnosis of learning disability or autism. As above, we used a Poisson ITSA model with robust standard errors to estimate the change in the subgroup, and the geometric mean of the relative risk comparing the model fitted values to the no COVID-19 counterfactual for an overall effect size. For all patients in either of these subgroups we calculated these measures where “any” and “new” antidepressants were prescribed.

#### Demographic and clinical subgroups

To estimate the impact of COVID-19 on demographic or clinical subgroups (age, sex, IMD decile, ethnicity, region, care home status, diagnosis, and antidepressant type), we created separate models for each subgroup (one model using only data for age 0-19 another model using only data for age 20-29 etc). As above, the relative risk comparing the model fitted value to the no COVID-19 counterfactual was computed for each time point, and then the geometric mean of those values was computed and those values were presented in a forest plot.

#### Sensitivity analysis

We also looked at total prescription counts, rather than number of patients. We first described the total rate of prescriptions issued in OpenSAFELY-TPP. Then we separately performed an ITSA with the same model and codelists, but using data from OpenPrescribing. OpenPrescribing is an openly available viewer of primary care prescription reimbursement data and contains information on prescription volumes at an aggregate practice level [31]. This data has been used by others to assess antidepressant prescribing [2,32].

## Results

### Population Characteristics

At the beginning of the study period in January 2018, there were 23,864,380 registered patients, increasing to 25,504,380 at the end of the study period in December 2022. In October 2022 (a representative month close to the end of the study period), 2,048,040 patients were prescribed an antidepressant, for an overall rate of prescribing of 80.5 per 1,000 registered patients (**Table 1**). The rate of prescribing was higher in women (107.0 per 1,000) than men (53.9 per 1,000). Prescribing increased with increasing IMD score (least deprived 69.7 versus most deprived 93.2 per 1,000) and increasing age (age group 0-19, 5.4 versus age group 80+, 139.2 per 1,000), and was highest in White ethnicity (98.5 per 1,000). Regionally, the North East (102.7 per 1,000) and North West (97.7 per 1,000) had higher prescribing than the other regions. Of patients aged 18+ and receiving an antidepressant prescription, 30% had neither an active depression diagnosis nor a record of anxiety.

**Table 1:** Characteristics of registered patients in October 2022 by total population, learning disability, or autism. Patients can be in both the learning disability and autism subgroups.

|  |  | All |  |  | Learning Disability |  |  | Autism |  |  |
| --- | --- | --- | --- | --- | --- | --- | --- | --- | --- | --- |
|  |  | No. prescribed antidepressant (%) | No. registered patients (%) | Rate per 1,000 (95% CI) | No. prescribed antidepressant (%) | No. registered patients (%) | Rate per 1,000 (95% CI) | No. prescribed antidepressant (%) | No. registered patients (%) | Rate per 1,000 (95% CI) |
| Total |  | 2048040 (100.0) | 25455570 (100.0) | 80.5 (80.3 to 80.6) | 26810 (100.0) | 145920 (100.0) | 183.7 (181.7 to 185.7) | 31360 (100.0) | 264610 (100.0) | 118.5 (117.3 to 119.7) |
| Age Band | 0-19 | 30200 (1.5) | 5582550 (21.9) | 5.4 (5.3 to 5.5) | 1060 (4.0) | 29370 (20.1) | 36.1 (34.0 to 38.2) | 5240 (16.7) | 152540 (57.7) | 34.4 (33.4 to 35.3) |
|  | 20-29 | 185450 (9.1) | 3130490 (12.3) | 59.2 (59.0 to 59.5) | 5210 (19.4) | 32690 (22.4) | 159.4 (155.4 to 163.3) | 12080 (38.5) | 64050 (24.2) | 188.6 (185.6 to 191.6) |
|  | 30-39 | 273300 (13.3) | 3671930 (14.4) | 74.4 (74.2 to 74.7) | 5460 (20.4) | 27030 (18.5) | 202.0 (197.2 to 206.8) | 6360 (20.3) | 25340 (9.6) | 251.0 (245.6 to 256.3) |
|  | 40-49 | 323540 (15.8) | 3269820 (12.8) | 98.9 (98.6 to 99.3) | 4270 (15.9) | 17660 (12.1) | 241.8 (235.5 to 248.1) | 3450 (11.0) | 10450 (3.9) | 330.1 (321.1 to 339.2) |
|  | 50-59 | 431840 (21.1) | 3458490 (13.6) | 124.9 (124.5 to 125.2) | 5330 (19.9) | 19380 (13.3) | 275.0 (268.7 to 281.3) | 2770 (8.8) | 7830 (3.0) | 353.8 (343.2 to 364.4) |
|  | 60-69 | 355050 (17.3) | 2816370 (11.1) | 126.1 (125.7 to 126.5) | 3690 (13.8) | 13020 (8.9) | 283.4 (275.7 to 291.2) | 1130 (3.6) | 3410 (1.3) | 331.4 (315.6 to 347.2) |
|  | 70-79 | 269310 (13.1) | 2237650 (8.8) | 120.4 (119.9 to 120.8) | 1500 (5.6) | 5510 (3.8) | 272.2 (260.5 to 284.0) | 280 (0.9) | 830 (0.3) | 337.3 (305.2 to 369.5) |
|  | 80+ | 179350 (8.8) | 1288270 (5.1) | 139.2 (138.6 to 139.8) | 300 (1.1) | 1240 (0.8) | 241.9 (218.1 to 265.8) | 50 (0.2) | 140 (<0.1) | 357.1 (277.8 to 436.5) |
| Carehome | False | 1982340 (96.8) | 25280560 (99.3) | 78.4 (78.3 to 78.5) | 19670 (73.4) | 122820 (84.2) | 160.2 (158.1 to 162.2) | 28930 (92.3) | 257480 (97.3) | 112.4 (111.1 to 113.6) |
|  | True | 65700 (3.2) | 175010 (0.7) | 375.4 (373.1 to 377.7) | 7140 (26.6) | 23100 (15.8) | 309.1 (303.1 to 315.1) | 2430 (7.7) | 7120 (2.7) | 341.3 (330.3 to 352.3) |
| Diagnosis (over 18s only) | Anxiety | 537670 (26.4) | 2636550 (12.9) | 203.9 (203.4 to 204.4) | 8020 (30.5) | 22260 (18.2) | 360.3 (354.0 to 366.6) | 9280 (32.3) | 30860 (23.6) | 300.7 (295.6 to 305.8) |
|  | Both | 598340 (29.4) | 1728650 (8.5) | 346.1 (345.4 to 346.8) | 5280 (20.1) | 10310 (8.4) | 512.1 (502.5 to 521.8) | 10200 (35.5) | 24550 (18.7) | 415.5 (409.3 to 421.6) |
|  | Depression Register | 287140 (14.1) | 1076850 (5.3) | 266.6 (265.8 to 267.5) | 3240 (12.3) | 6660 (5.4) | 486.5 (474.5 to 498.5) | 2960 (10.3) | 8580 (6.6) | 345.0 (334.9 to 355.0) |
|  | Neither | 614500 (30.2) | 14977500 (73.3) | 41.0 (40.9 to 41.1) | 9780 (37.2) | 83390 (68.0) | 117.3 (115.1 to 119.5) | 6330 (22.0) | 66980 (51.1) | 94.5 (92.3 to 96.7) |
| Ethnicity | Asian Or Asian British | 56840 (2.8) | 1823980 (7.2) | 31.2 (30.9 to 31.4) | 790 (2.9) | 8880 (6.1) | 89.0 (83.0 to 94.9) | 440 (1.4) | 8510 (3.2) | 51.7 (47.0 to 56.4) |
|  | ----Any Other Asian Background | 11560 (0.6) | 428410 (1.7) | 27.0 (26.5 to 27.5) | 130 (0.5) | 1460 (1.0) | 89.0 (74.4 to 103.7) | 100 (0.3) | 1840 (0.7) | 54.3 (44.0 to 64.7) |
|  | ----Bangladeshi | 4770 (0.2) | 127600 (0.5) | 37.4 (36.3 to 38.4) | 50 (0.2) | 780 (0.5) | 64.1 (46.9 to 81.3) | 30 (<0.1) | 1030 (0.4) | 29.1 (18.9 to 39.4) |
|  | ----Indian | 18770 (0.9) | 744810 (2.9) | 25.2 (24.8 to 25.6) | 220 (0.8) | 2250 (1.5) | 97.8 (85.5 to 110.1) | 160 (0.5) | 2350 (0.9) | 68.1 (57.9 to 78.3) |
|  | ----Pakistani | 21740 (1.1) | 523170 (2.1) | 41.6 (41.0 to 42.1) | 380 (1.4) | 4400 (3.0) | 86.4 (78.1 to 94.7) | 160 (0.5) | 3290 (1.2) | 48.6 (41.3 to 56.0) |
|  | Black Or Black British | 17600 (0.9) | 634380 (2.5) | 27.7 (27.3 to 28.1) | 260 (1.0) | 2900 (2.0) | 89.7 (79.3 to 100.1) | 210 (0.7) | 4430 (1.7) | 47.4 (41.1 to 53.7) |
|  | ----African | 7880 (0.4) | 411600 (1.6) | 19.1 (18.7 to 19.6) | 60 (0.2) | 1320 (0.9) | 45.5 (34.2 to 56.7) | 60 (0.2) | 2470 (0.9) | 24.3 (18.2 to 30.4) |
|  | ----Any Other Black Background | 3900 (0.2) | 106530 (0.4) | 36.6 (35.5 to 37.7) | 90 (0.3) | 740 (0.5) | 121.6 (98.1 to 145.2) | 70 (0.2) | 1060 (0.4) | 66.0 (51.1 to 81.0) |
|  | ----Caribbean | 5830 (0.3) | 116250 (0.5) | 50.2 (48.9 to 51.4) | 110 (0.4) | 850 (0.6) | 129.4 (106.8 to 152.0) | 80 (0.3) | 910 (0.3) | 87.9 (69.5 to 106.3) |
|  | Mixed | 15870 (0.8) | 387750 (1.5) | 40.9 (40.3 to 41.6) | 240 (0.9) | 2090 (1.4) | 114.8 (101.2 to 128.5) | 420 (1.3) | 5650 (2.1) | 74.3 (67.5 to 81.2) |
|  | ----Any Other Mixed Background | 5550 (0.3) | 144210 (0.6) | 38.5 (37.5 to 39.5) | 90 (0.3) | 760 (0.5) | 118.4 (95.4 to 141.4) | 160 (0.5) | 1990 (0.8) | 80.4 (68.5 to 92.3) |
|  | ----White And Asian | 3160 (0.2) | 83440 (0.3) | 37.9 (36.6 to 39.2) | 50 (0.2) | 390 (0.3) | 128.2 (95.0 to 161.4) | 80 (0.3) | 1070 (0.4) | 74.8 (59.0 to 90.5) |
|  | ----White And Black African | 2300 (0.1) | 74160 (0.3) | 31.0 (29.8 to 32.3) | 20 (<0.1) | 310 (0.2) | 64.5 (37.2 to 91.9) | 50 (0.2) | 860 (0.3) | 58.1 (42.5 to 73.8) |
|  | ----White And Black Caribbean | 4870 (0.2) | 85950 (0.3) | 56.7 (55.1 to 58.2) | 80 (0.3) | 630 (0.4) | 127.0 (101.0 to 153.0) | 130 (0.4) | 1720 (0.7) | 75.6 (63.1 to 88.1) |
|  | Other Ethnic Groups | 15250 (0.7) | 538860 (2.1) | 28.3 (27.9 to 28.7) | 130 (0.5) | 1110 (0.8) | 117.1 (98.2 to 136.0) | 160 (0.5) | 2240 (0.8) | 71.4 (60.8 to 82.1) |
|  | ----Any Other Ethnic Group | 13320 (0.7) | 343990 (1.4) | 38.7 (38.1 to 39.4) | 100 (0.4) | 900 (0.6) | 111.1 (90.6 to 131.6) | 120 (0.4) | 1670 (0.6) | 71.9 (59.5 to 84.2) |
|  | ----Chinese | 1930 (<0.1) | 194880 (0.8) | 9.9 (9.5 to 10.3) | 30 (0.1) | 220 (0.2) | 136.4 (91.0 to 181.7) | 40 (0.1) | 570 (0.2) | 70.2 (49.2 to 91.1) |
|  | White | 1660980 (81.1) | 16863830 (66.2) | 98.5 (98.4 to 98.6) | 22680 (84.6) | 109240 (74.9) | 207.6 (205.2 to 210.0) | 24230 (77.3) | 177810 (67.2) | 136.3 (134.7 to 137.9) |
|  | ----Any Other White Background | 111760 (5.5) | 2350800 (9.2) | 47.5 (47.3 to 47.8) | 1120 (4.2) | 6720 (4.6) | 166.7 (157.8 to 175.6) | 1310 (4.2) | 12070 (4.6) | 108.5 (103.0 to 114.1) |
|  | ----British | 1538070 (75.1) | 14400910 (56.6) | 106.8 (106.6 to 107.0) | 21470 (80.1) | 102100 (70.0) | 210.3 (207.8 to 212.8) | 22810 (72.7) | 165140 (62.4) | 138.1 (136.5 to 139.8) |
|  | ----Irish | 11150 (0.5) | 112130 (0.4) | 99.4 (97.7 to 101.2) | 100 (0.4) | 420 (0.3) | 238.1 (197.4 to 278.8) | 110 (0.4) | 600 (0.2) | 183.3 (152.4 to 214.3) |
|  | Missing | 281490 (13.7) | 5206760 (20.5) | 54.1 (53.9 to 54.3) | 2700 (10.1) | 21680 (14.9) | 124.5 (120.1 to 128.9) | 5890 (18.8) | 65960 (24.9) | 89.3 (87.1 to 91.5) |
| Imd | 1 - Most Deprived | 468560 (22.9) | 5025740 (19.7) | 93.2 (93.0 to 93.5) | 7960 (29.7) | 42570 (29.2) | 187.0 (183.3 to 190.7) | 7110 (22.7) | 65080 (24.6) | 109.3 (106.9 to 111.6) |
|  | 2 | 412720 (20.2) | 4912660 (19.3) | 84.0 (83.8 to 84.3) | 6320 (23.6) | 32880 (22.5) | 192.2 (188.0 to 196.5) | 6720 (21.4) | 55600 (21.0) | 120.9 (118.2 to 123.6) |
|  | 3 | 420100 (20.5) | 5224810 (20.5) | 80.4 (80.2 to 80.6) | 5470 (20.4) | 28830 (19.8) | 189.7 (185.2 to 194.3) | 6590 (21.0) | 52900 (20.0) | 124.6 (121.8 to 127.4) |
|  | 4 | 372570 (18.2) | 4909320 (19.3) | 75.9 (75.7 to 76.1) | 3920 (14.6) | 22200 (15.2) | 176.6 (171.6 to 181.6) | 5450 (17.4) | 44700 (16.9) | 121.9 (118.9 to 125.0) |
|  | 5 - Least Deprived | 315030 (15.4) | 4517350 (17.7) | 69.7 (69.5 to 70.0) | 2560 (9.5) | 16040 (11.0) | 159.6 (153.9 to 165.3) | 4560 (14.5) | 37540 (14.2) | 121.5 (118.2 to 124.8) |
|  | Missing | 59050 (2.9) | 865690 (3.4) | 68.2 (67.7 to 68.7) | 580 (2.2) | 3400 (2.3) | 170.6 (157.9 to 183.2) | 930 (3.0) | 8770 (3.3) | 106.0 (99.6 to 112.5) |
| Region | East | 478510 (23.4) | 5831400 (22.9) | 82.1 (81.8 to 82.3) | 6080 (22.7) | 30420 (20.8) | 199.9 (195.4 to 204.4) | 6920 (22.1) | 56400 (21.3) | 122.7 (120.0 to 125.4) |
|  | East Midlands | 391670 (19.1) | 4427330 (17.4) | 88.5 (88.2 to 88.7) | 4900 (18.3) | 25040 (17.2) | 195.7 (190.8 to 200.6) | 6380 (20.3) | 52560 (19.9) | 121.4 (118.6 to 124.2) |
|  | London | 55040 (2.7) | 1816040 (7.1) | 30.3 (30.1 to 30.6) | 520 (1.9) | 5710 (3.9) | 91.1 (83.6 to 98.5) | 620 (2.0) | 9930 (3.8) | 62.4 (57.7 to 67.2) |
|  | North East | 121680 (5.9) | 1184850 (4.7) | 102.7 (102.1 to 103.2) | 1620 (6.0) | 7950 (5.4) | 203.8 (194.9 to 212.6) | 2050 (6.5) | 16860 (6.4) | 121.6 (116.7 to 126.5) |
|  | North West | 215360 (10.5) | 2203770 (8.7) | 97.7 (97.3 to 98.1) | 2890 (10.8) | 15170 (10.4) | 190.5 (184.3 to 196.8) | 2980 (9.5) | 24660 (9.3) | 120.8 (116.8 to 124.9) |
|  | South East | 118880 (5.8) | 1654200 (6.5) | 71.9 (71.5 to 72.3) | 1750 (6.5) | 9500 (6.5) | 184.2 (176.4 to 192.0) | 2280 (7.3) | 18440 (7.0) | 123.6 (118.9 to 128.4) |
|  | South West | 289920 (14.2) | 3527600 (13.9) | 82.2 (81.9 to 82.5) | 4070 (15.2) | 21320 (14.6) | 190.9 (185.6 to 196.2) | 5400 (17.2) | 40970 (15.5) | 131.8 (128.5 to 135.1) |
|  | West Midlands | 72470 (3.5) | 1031020 (4.1) | 70.3 (69.8 to 70.8) | 1090 (4.1) | 6600 (4.5) | 165.2 (156.2 to 174.1) | 1210 (3.9) | 11840 (4.5) | 102.2 (96.7 to 107.7) |
|  | Yorkshire And The Humber | 297980 (14.5) | 3691560 (14.5) | 80.7 (80.4 to 81.0) | 3790 (14.1) | 23770 (16.3) | 159.4 (154.8 to 164.1) | 3420 (10.9) | 32160 (12.2) | 106.3 (103.0 to 109.7) |
|  | Missing | 6520 (0.3) | 87800 (0.3) | 74.3 (72.5 to 76.0) | 90 (0.3) | 440 (0.3) | 204.5 (166.9 to 242.2) | 110 (0.4) | 770 (0.3) | 142.9 (118.1 to 167.6) |
| Sex | F | 1360440 (66.4) | 12709460 (49.9) | 107.0 (106.9 to 107.2) | 13130 (49.0) | 57170 (39.2) | 229.7 (226.2 to 233.1) | 12850 (41.0) | 72380 (27.4) | 177.5 (174.8 to 180.3) |
|  | M | 687600 (33.6) | 12746120 (50.1) | 53.9 (53.8 to 54.1) | 13680 (51.0) | 88750 (60.8) | 154.1 (151.8 to 156.5) | 18510 (59.0) | 192230 (72.6) | 96.3 (95.0 to 97.6) |

IMD = index of multiple deprivation
Counts have been rounded to the nearest 10 and the rate computed with the rounded numbers
Group sums may not exactly match the "Total" due to rounding

In October 2022, there were 145,920 registered patients with recorded learning disability and 264,610 registered patients with recorded autism (patients could be counted in both groups). Rates of antidepressant prescribing were higher among those with learning disability (183.7 per 1,000) followed by autism (118.5 per 1,000). The crude rate of antidepressant use in the absence of a diagnosis of anxiety or depression for those aged 18+ was 117.3 per 1000 in those with learning disability and 94.5 per 1000 in autistic patients, compared to 41.0 per 1000 in the general population. However if we look at the relative proportion of each diagnosis only among those who received a prescription, 37% of those with learning disability had neither an active depression diagnosis nor a record of anxiety. For those with autism, that number was 22% (compared to 30% in the general population). Demographic prescribing trends remained similar to the total population, except for IMD where the correlation was less strong.

The most commonly prescribed antidepressant for the total population in October 2022 was SSRIs (54%), followed by tricyclics (18%) and other (20%) (**Table 2**). 9% of patients were prescribed more than one class of antidepressant. For both the learning disability and autism groups, there was a greater proportion of SSRIs prescribed (68%, 70%), but a smaller proportion of tricyclics (7%, 5%).

**Table 2:**
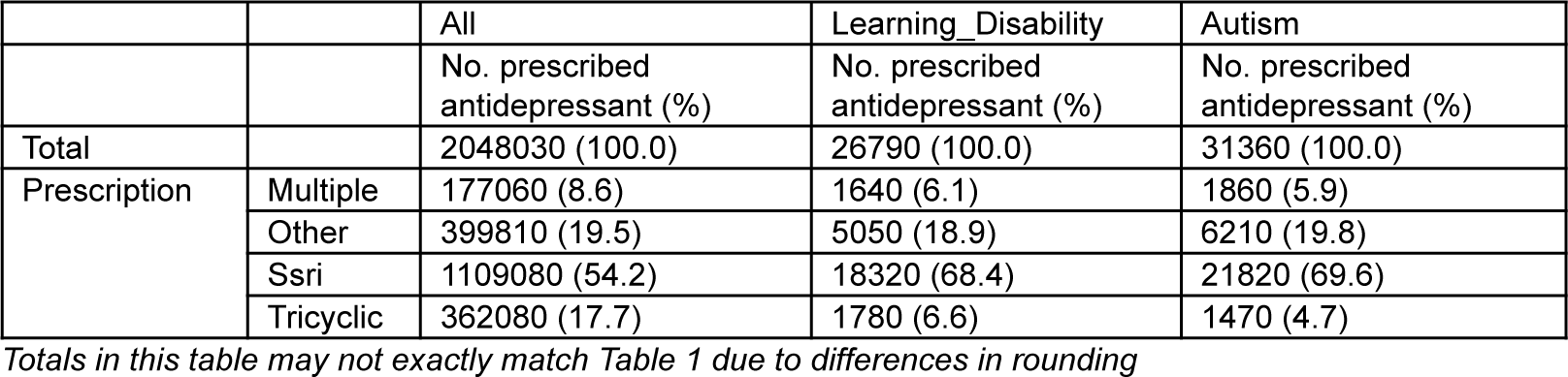
Relative proportion of each antidepressant class prescribed in October 2022 by total population, learning disability, or autism. Patients can be in both the learning disability and autism subgroups. Those prescribed more than one class of antidepressants in a single month are counted as ‘multiple’

### Trends in Overall Prescribing

Prior to the COVID-19 lockdown, antidepressant prescribing was increasing in the general population at a rate of 0.3% (95% CI 0.2% to 0.3%) per month (**Figure 1** and **Table 3**) (an increase in model fitted rate from 71.0 per 1,000 to 82.5 per 1,000).

**Figure 1:**
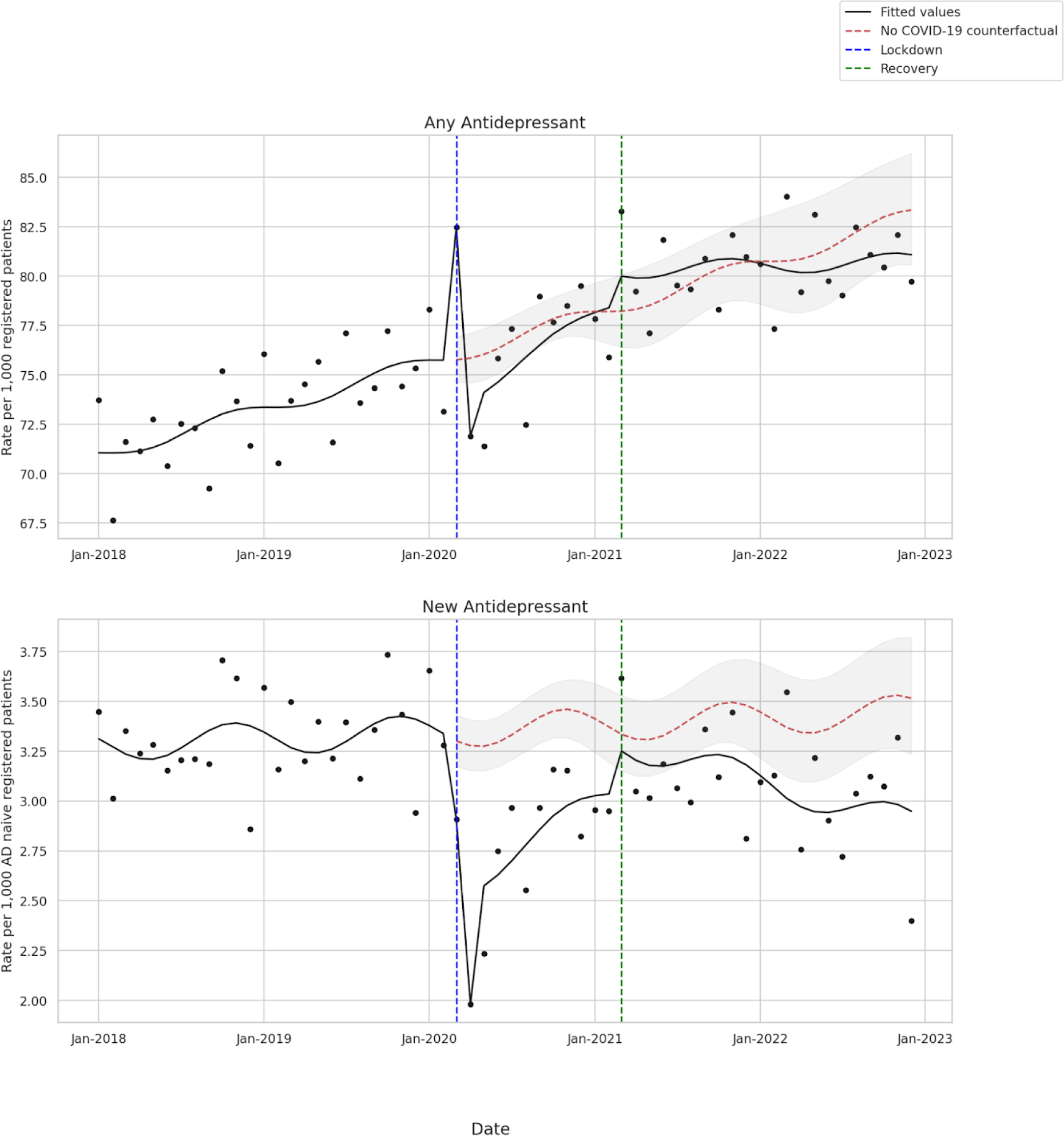
Rate per 1,000 of (top) antidepressant prescribing in the general population and (bottom) new antidepressant prescribing, adjusted for long-term seasonality and trend. The vertical dotted lines represent the start of the Lockdown period (March 2020 to February 2021) and the Recovery period (March 2021-December 2022). The dotted red line is the no COVID-19 counterfactual.

**Table 3:** Percent change in antidepressant prescribing during the Lockdown period (March 2020-February 2021) and Recovery period (March 2021-December 2022) adjusted for seasonality and long term trend.

|  |  | Lockdown period |  | Recovery period |  |
| --- | --- | --- | --- | --- | --- |
|  | Pre-COVID-19 monthly slope (95% CI) | Level shift (95% CI) | Change in slope (95% CI) | Level shift (95% CI) | Change in slope (95% CI) |
| All prescribing | 0.3% (0.2% to 0.3%) | -3.4% (-7.4% to 0.6%) | 0.3% (-0.2% to 0.9%) | 2.3% (-1.8% to 6.5%) | -0.5% (-1.0% to -0.1%) |
| Autism prescribing | 0.3% (0.2% to 0.4%) | -3.4% (-7.1% to 0.3%) | 0.4% (-0.1% to 0.9%) | 1.2% (-2.8% to 5.3%) | -0.7% (-1.2% to -0.3%) |
| LD prescribing | 0.3% (0.2% to 0.3%) | -2.9% (-5.4% to -0.4%) | 0.2% (-0.1% to 0.5%) | 0.8% (-1.8% to 3.4%) | -0.4% (-0.7% to -0.1%) |
| New prescribing | 0.1% (-0.1% to 0.3%) | -24.8% (-33.5% to -15.1%) | 1.5% (0.0% to 3.0%) | 9.1% (-0.3% to 19.4%) | -2.2% (-3.7% to -0.7%) |
| Autism new prescribing | 0.1% (-0.0% to 0.3%) | -21.7% (-32.8% to -8.9%) | 1.0% (-0.8% to 2.8%) | 5.8% (-3.9% to 16.5%) | -1.4% (-3.2% to 0.5%) |
| LD new prescribing | -0.0% (-0.3% to 0.3%) | -23.3% (-32.7% to -12.4%) | 2.8% (1.3% to 4.3%) | -8.4% (-18.5% to 3.0%) | -2.7% (-4.2% to -1.2%) |

Allowing for March and April to be outliers, there was no significant level shift or slope change with lockdown. Comparing the recovery period to lockdown, there was a negative slope change of −0.5% (95% CI −1.0% to −0.1%), but there was also no evidence of difference between the model fitted rate of antidepressant prescribing and the no COVID-19 counterfactual either in March 2021 (Recovery start) or in December 2022 (Study end) (**Figure S8**). Looking at the average post lockdown, we did not find evidence that the COVID-19 pandemic was associated with a change in antidepressant prescribing in the general population from the pre-COVID-19 trend (RR 1.00 (95% CI 0.97 to 1.02)) (**Figure 1** and **Figure S1**).

### Trends in New prescribing

Prior to the COVID-19 lockdown, the rate of new prescribing was stable (**Figure 1** and **Table 3**). At the beginning of lockdown there was a negative step change −24.8% (95% CI −33.5% to −−−−15.1%) but a non-significant slope change. However, comparing the recovery period to the lockdown period, there was a −2.2% (95% CI −3.7% to −0.7%) slope change.

At the start of the Recovery period in March 2021, there was no difference between the model fitted rate and the no COVID-19 counterfactual. At the end of the study period, however, there was −0.6 (95% CI −0.9 to −0.2) per 1,000 decrease in prescribing to antidepressant naive registered patients (**Figure S8**). The average relative risk of new prescribing post lockdown was 13% below expected if COVID-19 had not happened (RR 0.87 (95% CI 0.83 to 0.93)) (**Figure 1** and **Figure S1**).

### Autism and Learning Disability Subgroups

**Figure 2** and **Table 3** illustrate the impact of lockdown and recovery interruptions on prescribing of antidepressants in those with autism or learning disability.

**Figure 2:**
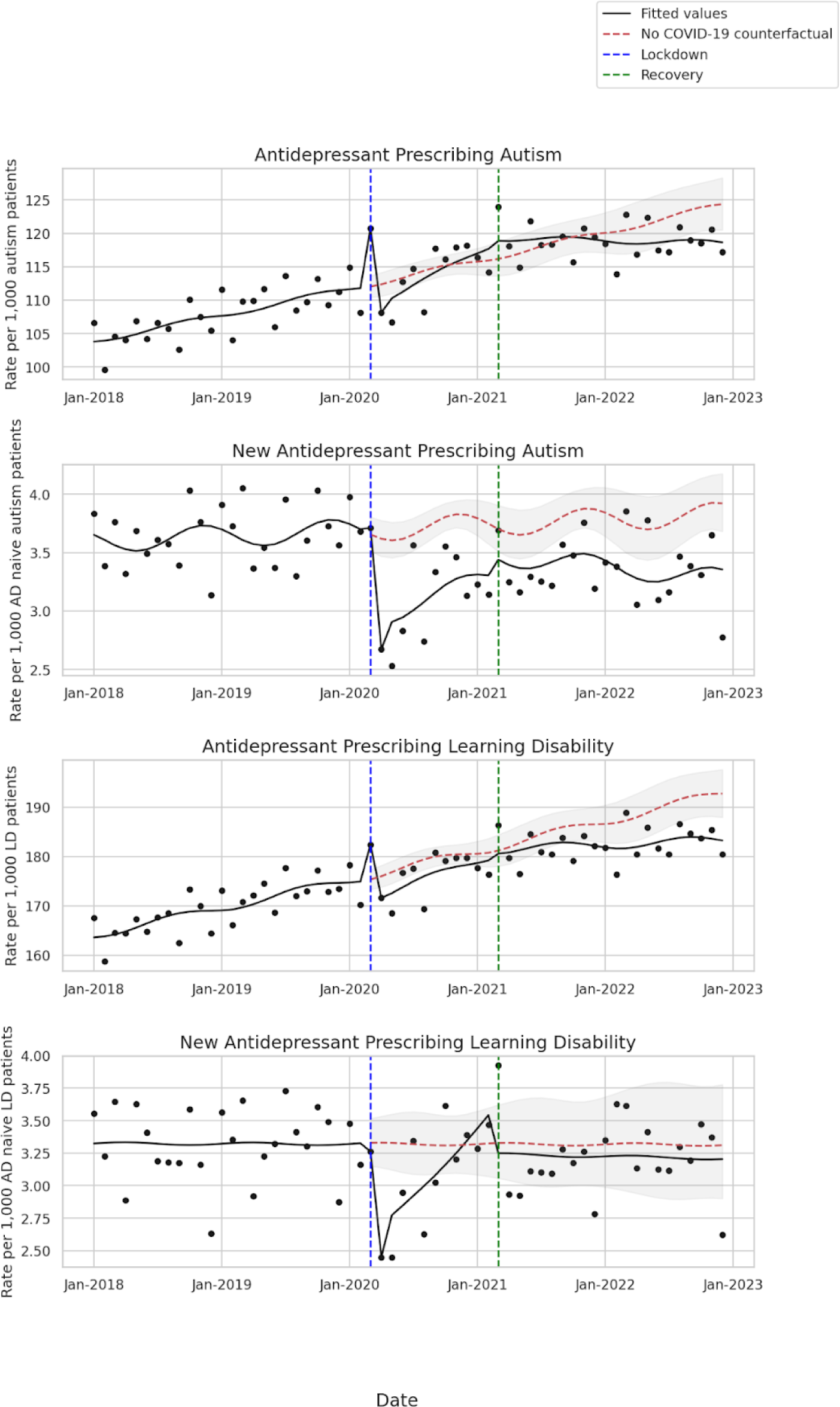
Predicted rate of patients prescribed an antidepressant per 1,000 registered patients among those with autism or learning disability from January 2018 to December 2022 adjusted for long-term seasonality and trend. The top row is among all autism or learning disability patients, and the second row is among those who have not been prescribed an antidepressant in the past 2 years (antidepressant naive). The vertical dotted lines represent the Lockdown period (March 2020 to February 2021) and the Recovery period (March 2021-December 2022). The dotted red line is the no COVID-19 counterfactual.

### Autism Overall Prescribing

In those with a record of autism, the findings were similar to the overall population. The baseline rate of any antidepressant prescribing was increasing 0.3% (95% CI 0.2% to 0.4%) per month (an increase in model fitted rate from 103.8 per 1,000 to 120.8 per 1,000). At the start of the Recovery, there was no difference between the model fitted rate and the no COVID-19 counterfactual, but in December 2022 the model fitted rate was −5.7 (95% CI −9.7 to −1.7) per 1,000 below the counterfactual (**Figure S8**). The average over the post lockdown period was not significantly different from the no COVID-19 counterfactual (RR 0.99 (95% CI 0.97 to 1.01)).

### Autism New Prescribing

In those with autism, new antidepressant prescribing was stable pre-COVID-19 and was 12% lower than expected had pre-COVID trends continued (RR 0.88 (95% CI 0.83 to 0.92)) (**Figure S2**). At the start of the Recovery, there was no difference between the model fitted rate of antidepressant prescribing and pre-COVID-19 trends, but in December 2022, the difference was −0.6 (95% CI −0.9 to −0.2) per 1,000 (**Figure S8**).

### Learning Disability Overall Prescribing

In those with a record of learning disability, the baseline rate of any antidepressant prescribing was also increasing 0.3% (95% CI 0.2% to 0.3%)) (an increase in model fitted rate from 163.6 to 182.4 per 1,000). As in the general and autism populations, the average over the post lockdown period was not significantly different from the no COVID-19 counterfactual (RR 0.98 (95% CI 0.96 to 1.00)). In absolute terms, there was no difference between the model fitted rate of antidepressant prescribing and the counterfactual (no COVID-19) at start of the Recovery, but in December 2022 the difference was −9.5 (95% CI −14.6 to −4.4) per 1,000 (**Figure S8**).

### Learning Disability New Prescribing

For new antidepressant prescribing to those with a record of learning disability, similar to the general population and those with autism, there was a negative level shift with Lockdown. Unlike in the general population or those with autism, however, there was a significant increase in slope during lockdown 2.8% (95% CI 1.3% to 4.3%). At the end of the study in December 2022, there was no evidence of any absolute difference from expected trends nor overall evidence that prescribing post Lockdown was lower than pre-COVID-19 trend (RR 0.96 (95% CI 0.87 to 1.05)) (**Figure S2**).

### Demographic and Clinical Subgroups

Although antidepressant prescribing in the general population post Lockdown was in line with pre-COVID-19 trends, it was 4% lower for under 30s compared to the no COVID-19 counterfactual (0-19 and 20-29 both RR 0.96 (95% CI 0.93 to 0.99)) (**Figure 3** and **Figure S5**). The pre-COVID-19 rate of patients prescribed tricyclics was decreasing at −0.1% (95% CI −0.2% to −0.1%), but post lockdown prescribing of tricyclics was 4% higher than expected (RR 1.04 95% CI 1.01 to 1.06) (**Figure 3** and **Figure S6**).

**Figure 3:**
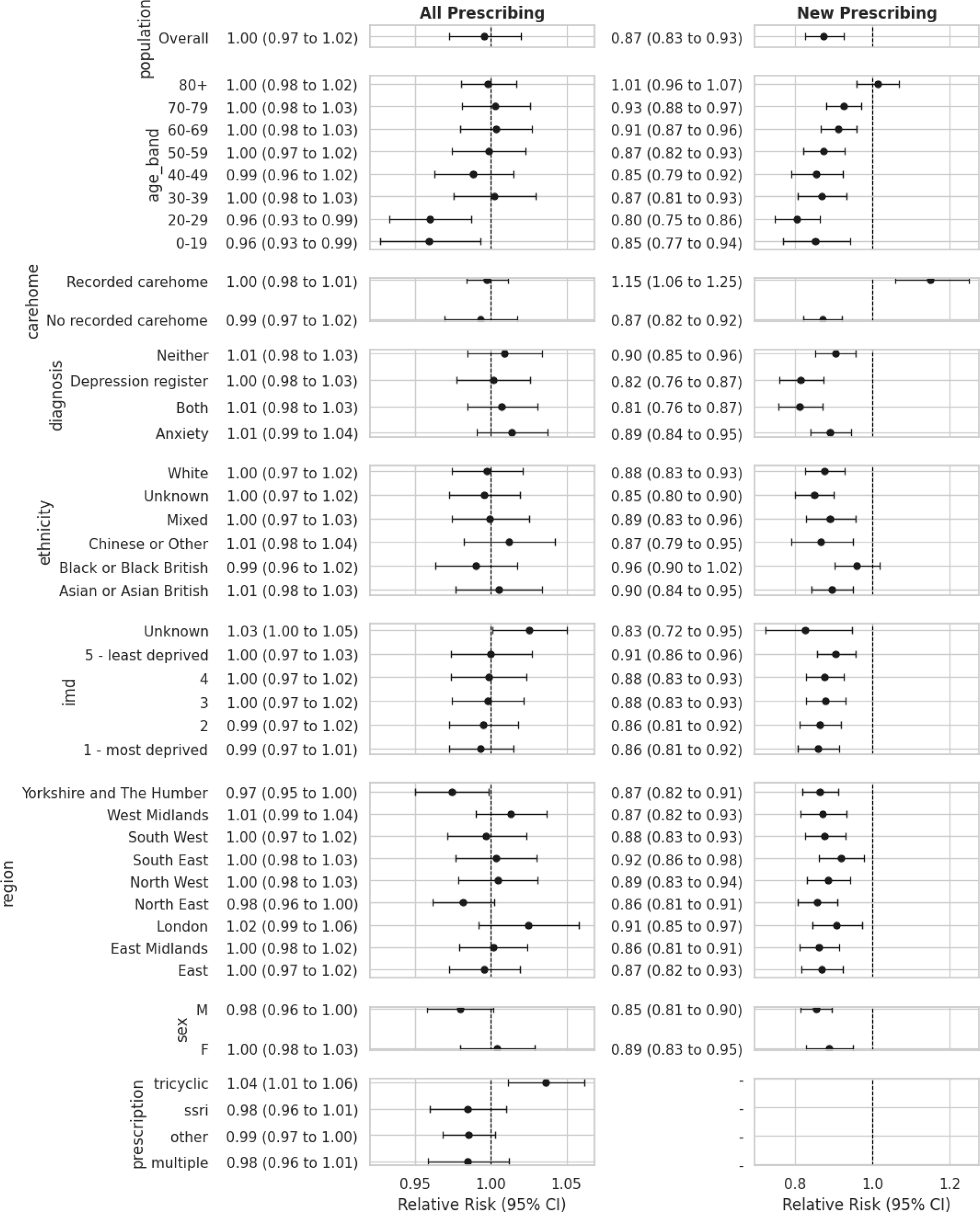
Overall and new antidepressant prescribing relative risks from March 2020 though December 2022 compared to the no COVID-19 counterfactual, by demographic subgroup. A relative risk of 1 represents no change from pre-COVID-19 trends.

New antidepressant prescribing in the general population was 13% lower than pre-COVID-19 trends would have predicted, and the difference with the counterfactual trend generally increased with decreasing age (80+ RR 1.01 (95% CI 0.95 to 1.07), versus 20-29 RR 0.80 (95% CI 0.75 to 0.86)) (**Figure 3**).

For those in a care home there was a 15% increase in new prescribing (**Figure 3**). In the final month of the study period, this meant a difference of 1.8 (95% CI 0.7 to 2.9) per 1,000 antidepressant naive patients (**Figure S9**).

There was no evidence that COVID was associated with a difference from pre-COVID-19 trends in overall antidepressant prescribing for any of the diagnosis groups (depression, anxiety, both, neither). For patients newly prescribed an antidepressant, the decrease from pre-COVID-19 trends was most pronounced in those on the depression register (with or without anxiety) compared to those with only anxiety or neither (**Figure 3, Figure S9**).

### Sensitivity analysis

Assessing trends using dispensing data in OpenPrescribing shows an increase in the rate of overall antidepressant prescribing from 96 per 1,000 in September 2018 to 117 per 1,000 in December 2022 (data in OpenPrescribing was available starting in September 2018). This was higher than the monthly rate of patients prescribed an antidepressant observed in this study (increasing from 69 per 1,000 to 80 per 1,000), indicating multiple antidepressant prescriptions may be collected per patient per month. An ITSA with OpenPrescribing data (**Figure S7**) showed that prescriptions dispensed and claimed for reimbursement in the general population were in line with what would have been expected had COVID-19 not happened, in line with our main results. When the total number of prescribing events (rather than the number of patients) was analysed using OpenSAFELY, the rates were slightly higher than OpenPrescribing (105 per 1,000 to 127 per 1,000). This is expected as not all prescriptions are collected by patients.

## Discussion

### Statement of principal findings

We did not see evidence that the COVID-19 pandemic had a sustained impact upon pre-COVID-19 antidepressant prescribing trends in the general population. At the start of the Recovery in March 2021, it appeared new prescribing to those without a record of a prescription in the past two years had recovered to the no COVID-19 counterfactual. After March 2021, however, the rate began to drop again and in the last month of the study period our model fitted rate was 0.5 (95% CI 0.2 to 0.9) per 1,000 lower than historical rates would have predicted. Overall, there appeared to be an average 13% decrease in new antidepressant prescribing. We are unable to tell, however, whether this is due to the COVID-19 pandemic, the impact of ongoing deprescribing initiatives, or some other cause.

Overall and new prescribing trends to those with autism were similar to those in the general population. For those with learning disability, however, the findings were different from the general population and autism findings: we did not see evidence that new prescribing was below expected trends, but overall prescribing was 2% lower than expected. New prescribing to those in a care home increased 15% from pre-COVID trends, and overall prescribing of tricyclics increased 3%. There was some evidence that overall prescribing decreased in 0-19 and 20-29 year olds, but it is important to note that prescribing to 0-19 year olds is often done in coordination with the Child and Adolescent Mental Health Services (CAMHS), a secondary care service.

There was no evidence that COVID was associated with a difference from pre-COVID-19 trends in overall antidepressant prescribing for any of the diagnosis groups (depression, anxiety, both, neither). In October 2022, our data shows that there were higher rates of diagnosis of depression and anxiety in those with learning disability and autism than the general population, but also that there were differences in the rates of prescribing without an indication between these populations. Of the adult patients receiving an antidepressant prescription, 30% in the general population had neither an active depression diagnosis nor a record of anxiety. That proportion was 37% among those with learning disability, and 22% for those with autism.

### Findings in context

A number of studies globally used ITSA to assess the impact of COVID-19 on mental health. Campitelli et al found a 1.43% increase in individuals dispensed antidepressants in nursing homes in Canada [33]. Estrela et al found a decreasing trend in daily dose of antidepressant prescribing for men in Portugal [34]. In Israel Frangou et al found COVID-19 was associated with an increase in antidepressant prescription fills [30]. Using data from the IQVIA National Prescription Audit in the US, Chai et al found no significant change in trend for new antidepressant prescriptions through March 2022 [35]. Wolfschlang et al found no observed change in psychotropic or antidepressant medications in one Swedish region [36]. The authors note that unlike other European countries, Sweden did not experience a true “lockdown”, highlighting that the impact of COVID-19 lockdowns may have been country-specific.

Using the UK Clinical Practice Research Datalink (CPRD) in England, Mansfield et al found substantial reduction in depression, anxiety, and self-harm primary care contacts in March 2020 that had not recovered by July 2020 [37]. We also saw that antidepressant prescribing and new antidepressant prescribing were below expected trends in July 2020, but allowing more follow-up time, we found that both had recovered by March 2021 (though new prescribing then again decreased, and on average was below expected trends).

Also using CPRD, Carr et al performed an ITSA of mental illness and self-harm, including antidepressant prescribing in England through September 10, 2020 [38]. In April 2020, they found a 36.4% decrease in first antidepressant prescriptions that had recovered by September 2020. Between March 1st and September 10th 2020, there was a 17.3% (95% CI 14.1 to 20.5) reduction in expected first antidepressant prescriptions. We saw a level shift of −29.8% for new prescriptions with lockdown, but we did not see new prescribing recover (before once again dropping) until later, in January or February of 2021. Our “new prescribing” outcome, however, includes those who had a previous diagnosis 2 or more years ago. This might be reflected in our higher baseline rate of new prescriptions (3.3 per 1,000 versus 2.2 per 1,000 in their study). Future research could look separately at the impact of COVID on those with a prescription from 2 or more years ago.

We did not see other ITSA analyses in England looking at the impact of COVID-19 on antidepressant prescribing or new initiation of antidepressants in at-risk groups such as those in care homes, or with learning disability or autism. However, Macdonald et al used OpenSAFELY to assess antipsychotic prescribing to those in care homes, and with learning disability or autism. Comparing Q1 2019 to Q4 2021, they found decreased antipsychotic prescribing to those with learning disability or autism, and an increase in new prescriptions to those in care homes [17], in accordance with our findings for antidepressants.

Since 2019, NHS England has maintained indicators to monitor antidepressant prescribing to those with learning disability and we can roughly validate our numbers against theirs. Branford et al [39] reported on these indicators and found 10.3% annual prevalence (calculated as number of patients with a prescription in the last 6 months) of antidepressant prescribing in the general population and 20.7% in those with learning disability for NHS financial year 2020-2021. When we calculated prevalence in the last 6 months of NHS financial year 2021 with our data, we found prevalence rates of 12.2% and 22.8%, respectively. Small differences between analyses are normal and expected due to differences in underlying populations.

### Strengths and limitations

A key strength of this paper is its scale; using the OpenSAFELY platform we have been able to access raw, pseudonymised, single-event-level clinical events for > 24 million patients in England, who are registered at NHS GP practices, that use TPP software. This allowed us to explore medication usage, diagnostic events, and other salient clinical, regional and demographic information including ethnicity, age and scores of deprivation.

There are, however, limitations to note. ITSA is a strong quasi-experimental study design that can help address confounding by using each population as its own control. But this uncontrolled ITSA cannot address ceiling effects or competing risks. A ceiling effect is the idea that there is a maximum number of patients who could be prescribed an antidepressant. Greater awareness around inappropriate prescribing, e.g. NICE guidance that antidepressants should not be offered as the first-line treatment for mild depression [33], is an example of a competing factor that could have impacted prescribing at the same time as the COVID-19 pandemic. A future controlled ITSA comparing those with severe versus non-severe depression could potentially help contextualise this effect.

This research relies on accurate recording of diagnosis, clinical events and interventions within primary care electronic notes. We are aware that conditions, such as learning disability are not well coded within primary care records and there are national incentives underway to improve this [40] but this is a limitation of all large EHR database research projects.

We see more differences from the no COVID-19 counterfactual in new prescribing than overall prescribing. New prescribing will be a mix of people completely new to antidepressants and people who have taken them previously, but more than 2 years ago. The pandemic may have affected these groups differently.

### Policy Implications and Interpretation

Following a Public Health England (PHE) review on medicines that may cause dependence or withdrawal [5], in March 2022 NICE published guidance [41] highlighting the need for a full and careful assessment, before prescribing antidepressants, especially in those with learning disability and autism [42]. Later that year, NICE launched their updated guidance on Depression, further stating the need to consider non-pharmacological options before using antidepressants [42].These guidance updates are in keeping with the trends that we observed towards decreased initiation of antidepressants across the population after the pandemic.

In 2021, a national review by the Chief Pharmaceutical Officer estimated that up to 10% of all medicines prescribed within the UK are no longer needed or not appropriate for continued use [43]. Notably, within this review, the authors highlighted a systemic problem of medicines being issued without a documented indication. Our findings suggest that up to one third of patients issued an antidepressant do not have a diagnosis of depression or anxiety recorded within their notes and this problem is even more prevalent in those with a diagnosis of learning disability. It is possible that GPs may have recorded other indications for some of these antidepressant prescriptions or recorded indications in freetext. Yet, our data supports the recommendations of the Chief Pharmaceutical Officer that improvements are needed in the documentation of diagnosis when prescribing medicines.

The key sentiments about medication review, optimisation and, where necessary, deprescribing agree with the principles of the STOMP initiative which was launched in 2016 in the UK [6]. Its aim was to reduce overprescribing of psychotropic medicines within the learning disability and autism populations, yet there is still work needed to improve prescribing practice within these groups. We found that patients with a learning disability prescribed an antidepressant are less likely to have a diagnosis for depression and/or anxiety recorded than the general population. Furthermore, we saw a significant increase in new antidepressant prescribing during the pandemic in patients that live in care homes, a particularly vulnerable group that includes those with a learning disability. We feel that more is needed to improve prescribing practice and ensure that antidepressants are used appropriately within these vulnerable populations.

We have shown how we can use the OpenSAFELY platform as a tool to monitor the impact of directives at a detailed and comprehensive level within ‘at-risk’ patient populations across the UK. Using the OpenSAFELY framework we can conduct rapid near real-time research into prescribing trends of medicines including antidepressants for almost the entire population of England. We can then focus in detail over a range of key variables, including medication type, diagnosis, demographics and ethnicity. With appropriate permissions and where appropriate support can be obtained from relevant professional bodies, the OpenSAFELY platform is also technically capable of providing audit and feedback information about clinical practice, and changes in clinical practice, at single sites to support improvements in patient care.

## Conclusion

We found that prior to the COVID-19 pandemic, antidepressant prescribing was increasing at about 0.3% per month. While we did not see an impact of the COVID-19 pandemic on overall prescribing in the general population, prescriptions to those aged 0-19, 20-29, and new prescriptions were lower than pre-COVID-19 trends would have predicted, but prescribing of tricyclics and new prescriptions to those in a care home were higher than expected.

## Data Availability

The dataset analysed within OpenSAFELY is based on > 24 million people currently registered with GP surgeries using TPP SystmOne software. All data were linked, stored and analysed securely using the OpenSAFELY platform, https://www.opensafely.org/, as part of the NHS England OpenSAFELY COVID-19 service. Data include pseudonymised data such as coded diagnoses, medications and physiological parameters. No free text data are included. All code is shared openly for review and re-use under MIT open licence. Detailed pseudonymised patient data is potentially re-identifiable and therefore not shared. Data management and analysis was performed using Python 3. All code for data management and analysis, as well as codelists, is shared openly for inspection and re-use.

https://github.com/opensafely/antidepressant-prescribing-lda

## Acknowledgements

We are very grateful for all the support received from the TPP Technical Operations team throughout this work, and for generous assistance from the information governance and database teams at NHS England and the NHS England Transformation Directorate.

## Information governance and ethical approval

NHS England is the data controller of the NHS England OpenSAFELY COVID-19 Service; TPP is the data processor; all study authors using OpenSAFELY have the approval of NHS England [44]. This implementation of OpenSAFELY is hosted within the TPP environment which is accredited to the ISO 27001 information security standard and is NHS IG Toolkit compliant [45].

Patient data has been pseudonymised for analysis and linkage using industry standard cryptographic hashing techniques; all pseudonymised datasets transmitted for linkage onto OpenSAFELY are encrypted; access to the NHS England OpenSAFELY COVID-19 service is via a virtual private network (VPN) connection; the researchers hold contracts with NHS England and only access the platform to initiate database queries and statistical models; all database activity is logged; only aggregate statistical outputs leave the platform environment following best practice for anonymisation of results such as statistical disclosure control for low cell counts [46].

The service adheres to the obligations of the UK General Data Protection Regulation (UK GDPR) and the Data Protection Act 2018. The service previously operated under notices initially issued in February 2020 by the the Secretary of State under Regulation 3(4) of the Health Service (Control of Patient Information) Regulations 2002 (COPI Regulations), which required organisations to process confidential patient information for COVID-19 purposes; this set aside the requirement for patient consent [47]. As of 1 July 2023, the Secretary of State has requested that NHS England continue to operate the Service under the COVID-19 Directions 2020 [48]. In some cases of data sharing, the common law duty of confidence is met using, for example, patient consent or support from the Health Research Authority Confidentiality Advisory Group [49].

Taken together, these provide the legal bases to link patient datasets using the service. GP practices, which provide access to the primary care data, are required to share relevant health information to support the public health response to the pandemic, and have been informed of how the service operates.

This study was approved by the Health Research Authority (REC reference 20/LO/0651).

## Guarantor

BMK is guarantor.

## Contributorship

All authors contributed to and approved the final manuscript.

## Abbreviations

BNF, EHR, IMD, ITSA, MAOI, QOF, RR, SSRI, STOMP

