## Supplementary Material for "The impact of the COVID-19 pandemic on Antidepressant Prescribing with a focus on people with learning disability and autism: An interrupted time-series analysis in England using OpenSAFELY-TPP"

### Antidepressant Prescribing - Supplementary Material

**Table S2.** Counts and relative percentages of registered patients excluded from the study in October 2022 by total population, learning disability, or autism. Patients can be in both the learning disability and autism subgroups. Counts <=5 are redacted. The next smallest group is also redacted to avoid potential re-identification of individuals. Numbers are rounded to the nearest 10 and percentages re-computed after redaction and rounding.

| | **All** | | **Learning Disability** | | **Autism** | |
|---|---|---|---|---|---|---|
| | No. prescribed an antidepressant (%) | No. registered patients (%) | No. prescribed an antidepressant (%) | No. registered patients (%) | No. prescribed an antidepressant (%) | No. registered patients (%) |
| Age Band | [REDACTED] | 10 (<0.01) | [REDACTED] | [REDACTED] | [REDACTED] | [REDACTED] |
| Sex | 180 (<0.01) | 1050 (<0.01) | [REDACTED] | [REDACTED] | 30 (<0.1) | 120 (<0.1) |

**Table S1.** Codelists used in this study

| **Codelist type** | **Codelist description** | **Codelists used in this study** | **OpenCodelists Reference Codelist*** |
|---|---|---|---|
| Antidepressants | Monoamine oxidase inhibitors | https://github.com/opensafely/antidepressant-prescribing-lda/blob/main/local_codelists/opensafely-monoamine-oxidase-inhibitors-dmd_new.csv | https://www.opencodelists.org/codelist/opensafely/monoamine-oxidase-inhibitors-dmd/2b785e16/ |
| | Tricyclic and related | https://github.com/opensafely/antidepressant-prescribing-lda/blob/main/local_codelists/opensafely-tricyclic-and-related-antidepressants-dmd_new.csv | https://www.opencodelists.org/codelist/opensafely/tricyclic-and-related-antidepressants-dmd/1268eae1/ |
| | Selective serotonin reuptake inhibitors | https://github.com/opensafely/antidepressant-prescribing-lda/blob/main/local_codelists/opensafely-selective-serotonin-reuptake-inhibitors-dmd_new.csv | https://www.opencodelists.org/codelist/opensafely/selective-serotonin-reuptake-inhibitors-dmd/6a572e55/ |
| | Other antidepressants | https://github.com/opensafely/antidepressant-prescribing-lda/blob/main/local_codelists/opensafely-other-antidepressants-dmd_new.csv | https://www.opencodelists.org/codelist/opensafely/other-antidepressants-dmd/795977ab/ |
| Clinical Subgroups | Autism | https://github.com/opensafely/antidepressant-prescribing-lda/blob/main/codelists/nhsd-primary-care-domain-refsets-autism_cod.csv | https://www.opencodelists.org/codelist/nhsd-primary-care-domain-refsets/autism_cod/20210127/ |
| | Learning Disability | https://github.com/opensafely/antidepressant-prescribing-lda/blob/main/codelists/nhsd-primary-care-domain-refsets-ld_cod.csv | https://www.opencodelists.org/codelist/nhsd-primary-care-domain-refsets/ld_cod/20210127/ |
| | Anxiety | https://github.com/opensafely/antidepressant-prescribing-lda/blob/main/codelists/opensafely-anxiety-disorders.csv | https://www.opencodelists.org/codelist/opensafely/anxiety-disorders/6aef605a/ |

* Codelists used in the study may differ from the reference codelists on OpenCodelists due to updates in dm+d codes representing specific medicines. Codes used in this study were up-to-date at the time of data analysis. See the documentation on keeping codelists up to date for more details.

**Figure S1.** Relative risk of antidepressant prescribing in the general population comparing the model fitted rate compared to the no COVID-19 counterfactual from March 2020 though December 2022. A relative risk of 1 represents no change from pre-COVID-19 trends. The overall effect size is computed with the geometric mean of all the time points.

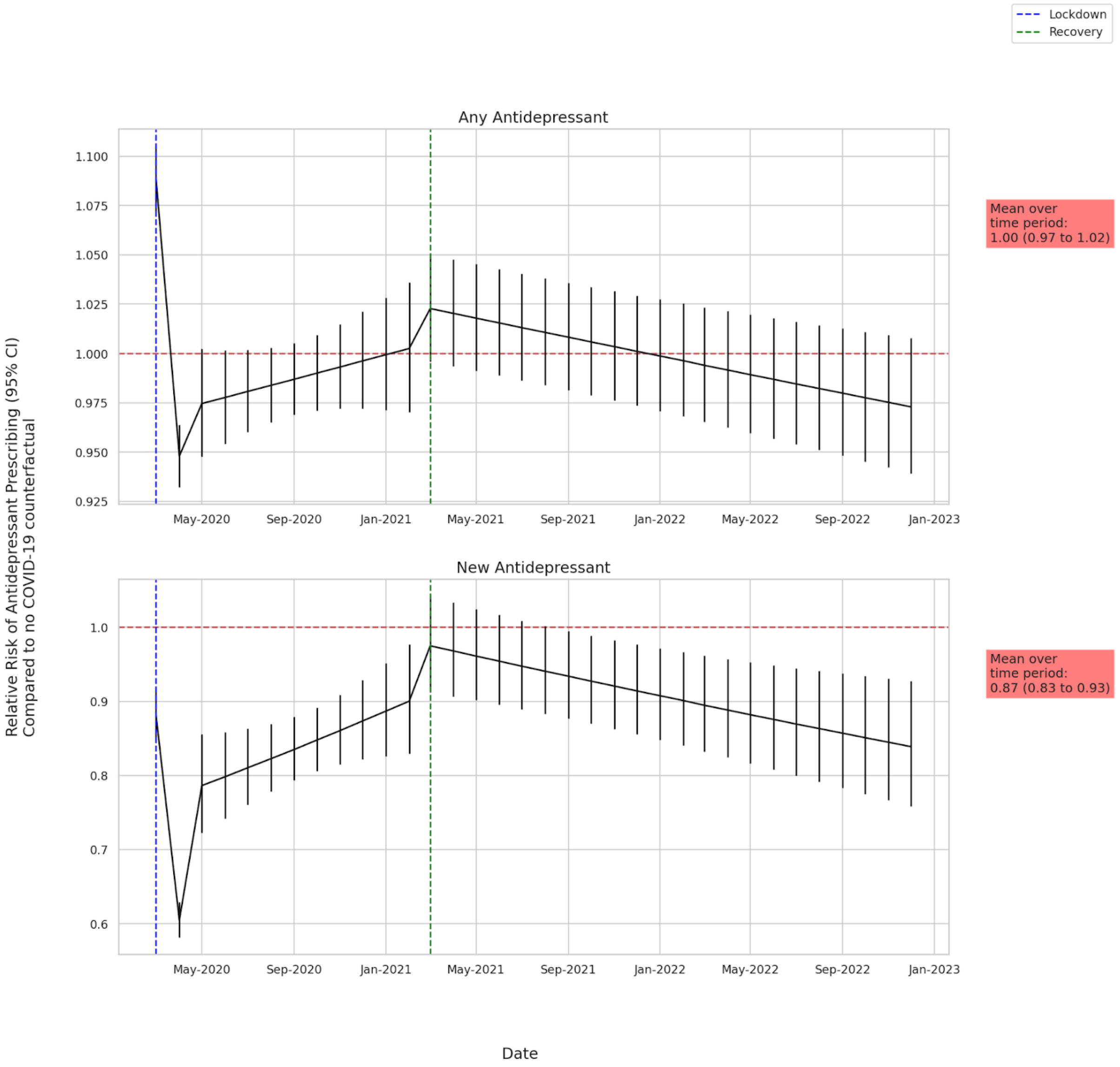

**Figure S2:** Relative risk of antidepressant prescribing for those with autism or learning disability comparing the model fitted rate compared to the no COVID-19 counterfactual from March 2020 though December 2022. A relative risk of 1 represents no change from pre-COVID-19 trends. The overall effect size is computed with the geometric mean of all the time points.

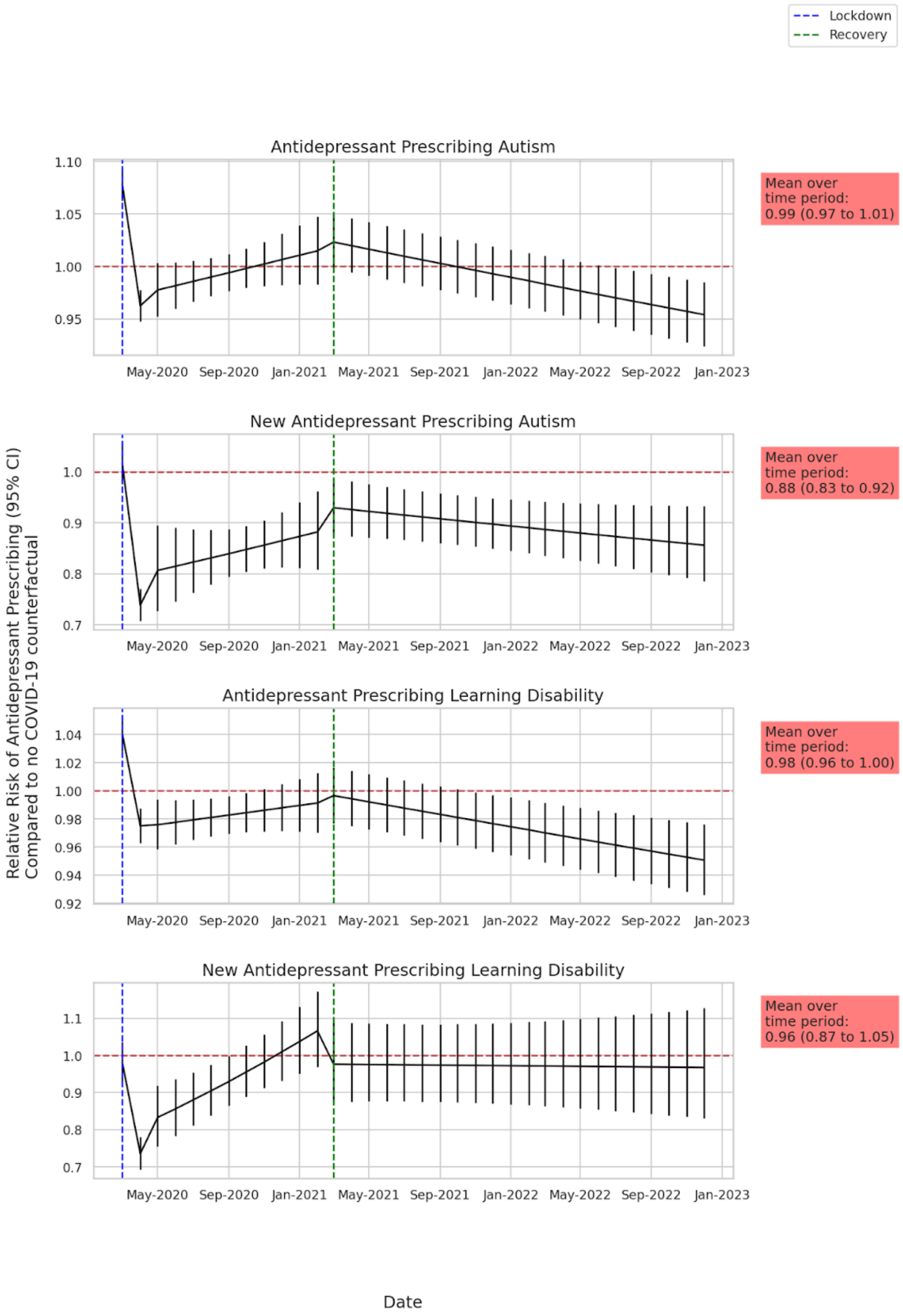

**Figure S3.**: Predicted rate of patients prescribed an antidepressant per 1,000 registered patients from January 2018 to December 2022 by demographic group. Each demographic subgroup is modeled separately.

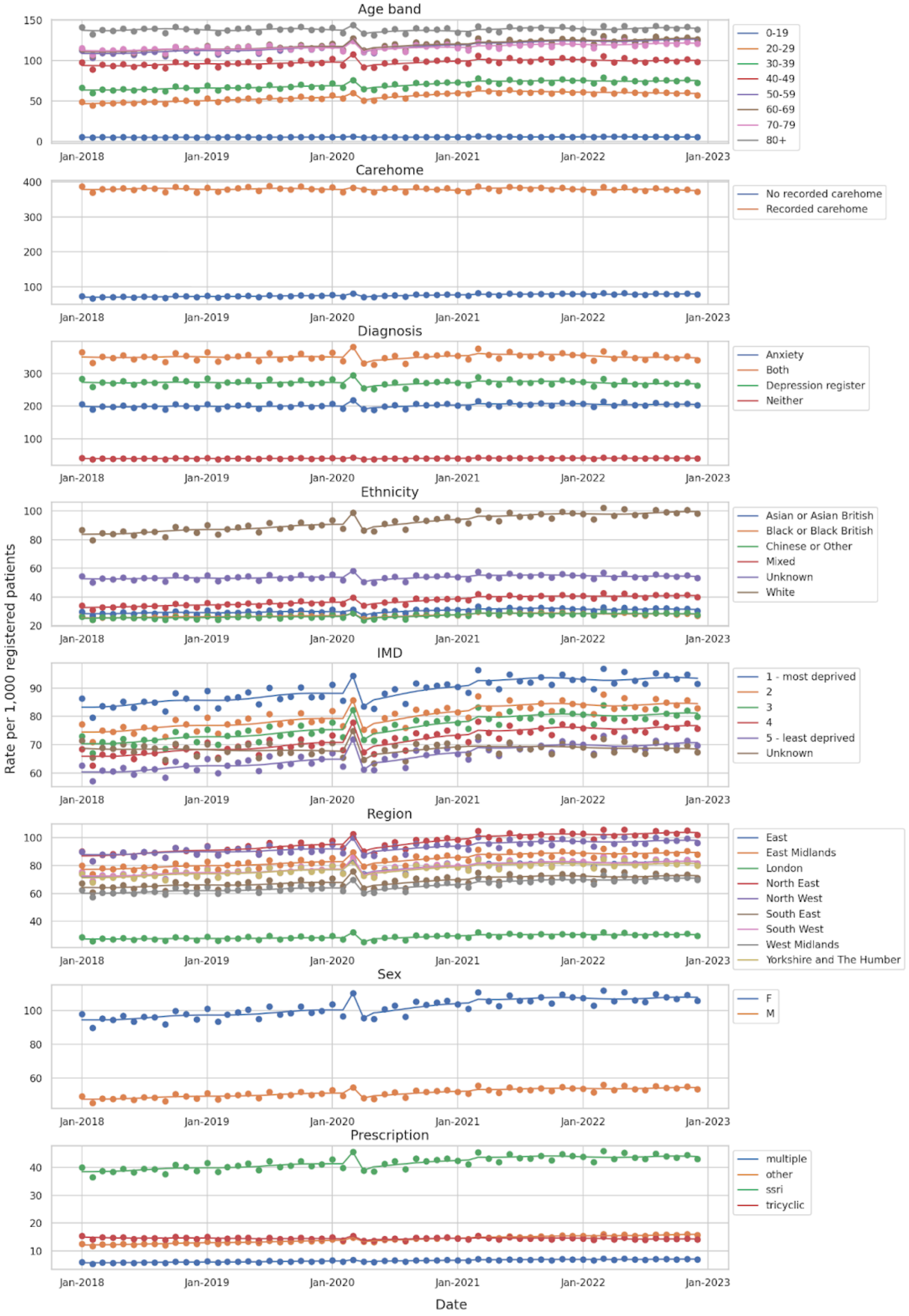

**Figure S4:** Predicted rate of patients prescribed an antidepressant per 1,000 registered patients 18 years and older in the general population, broken down by diagnosis from January 2018 to December 2022 adjusted for long-term seasonality and trend. The vertical dotted lines represent the Lockdown period (March 2020 to February 2021) and the Recovery period (March 2021-December 2022). The dotted red line is the no COVID-19 counterfactual.

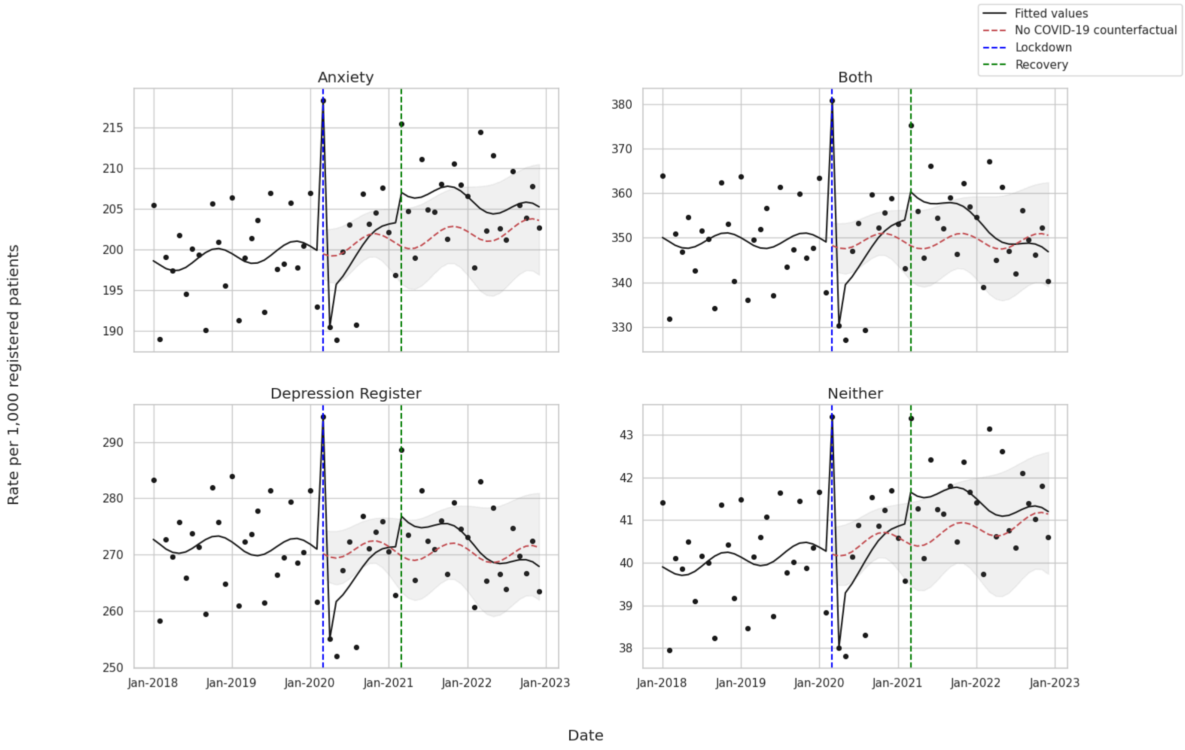

**Figure S5:** Predicted rate of patients prescribed an antidepressant per 1,000 registered patients in the general population, broken down by age band from January 2018 to December 2022 adjusted for long-term seasonality and trend. The vertical dotted lines represent the Lockdown period (March 2020 to February 2021) and the Recovery period (March 2021-December 2022). The dotted red line is the no COVID-19 counterfactual.

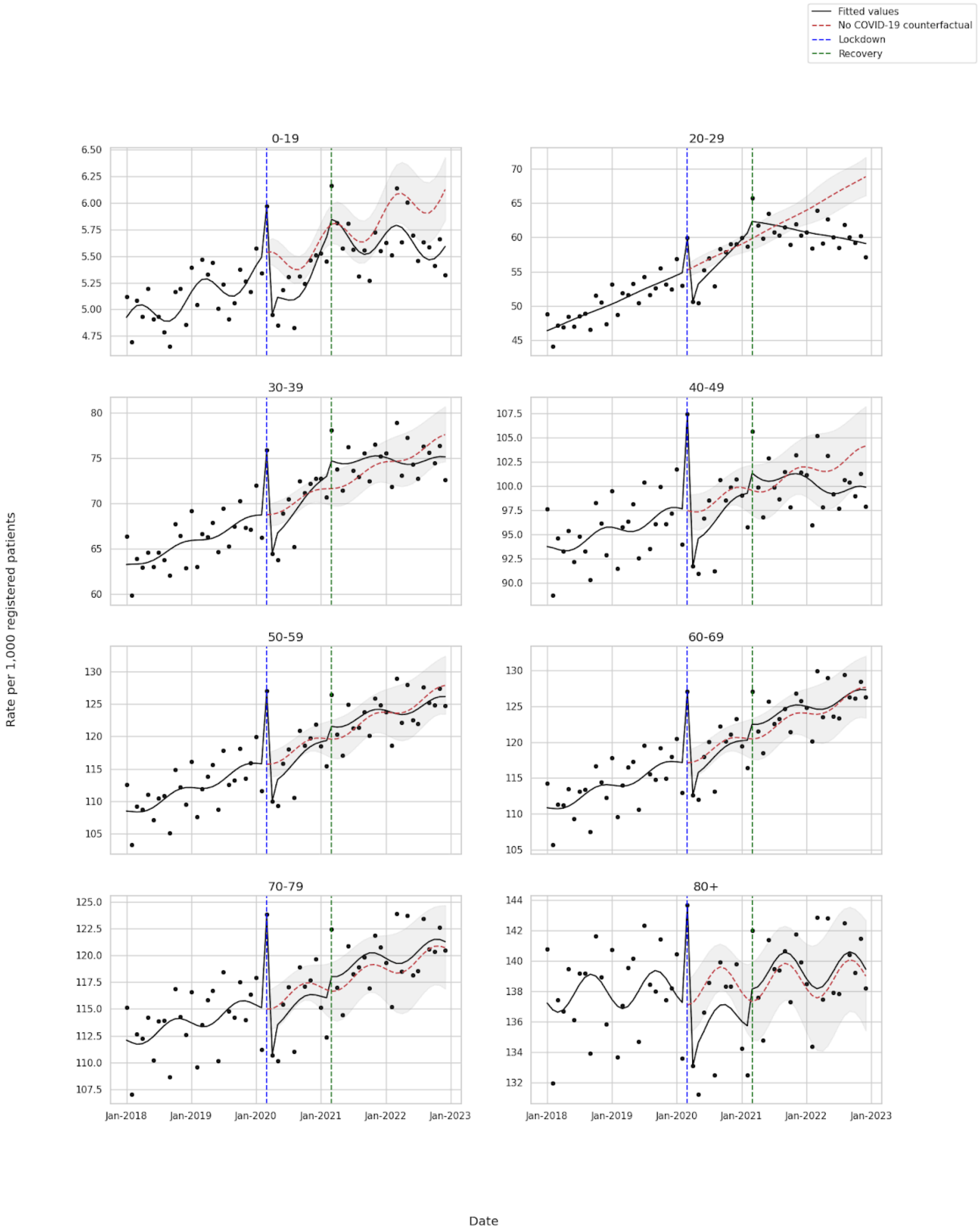

**Figure S6:** Predicted rate of patients prescribed an antidepressant per 1,000 registered patients in the general population, broken down by prescription type from January 2018 to December 2022 adjusted for long-term seasonality and trend. The vertical dotted lines represent the Lockdown period (March 2020 to February 2021) and the Recovery period (March 2021-December 2022). The dotted red line is the no COVID-19 counterfactual.

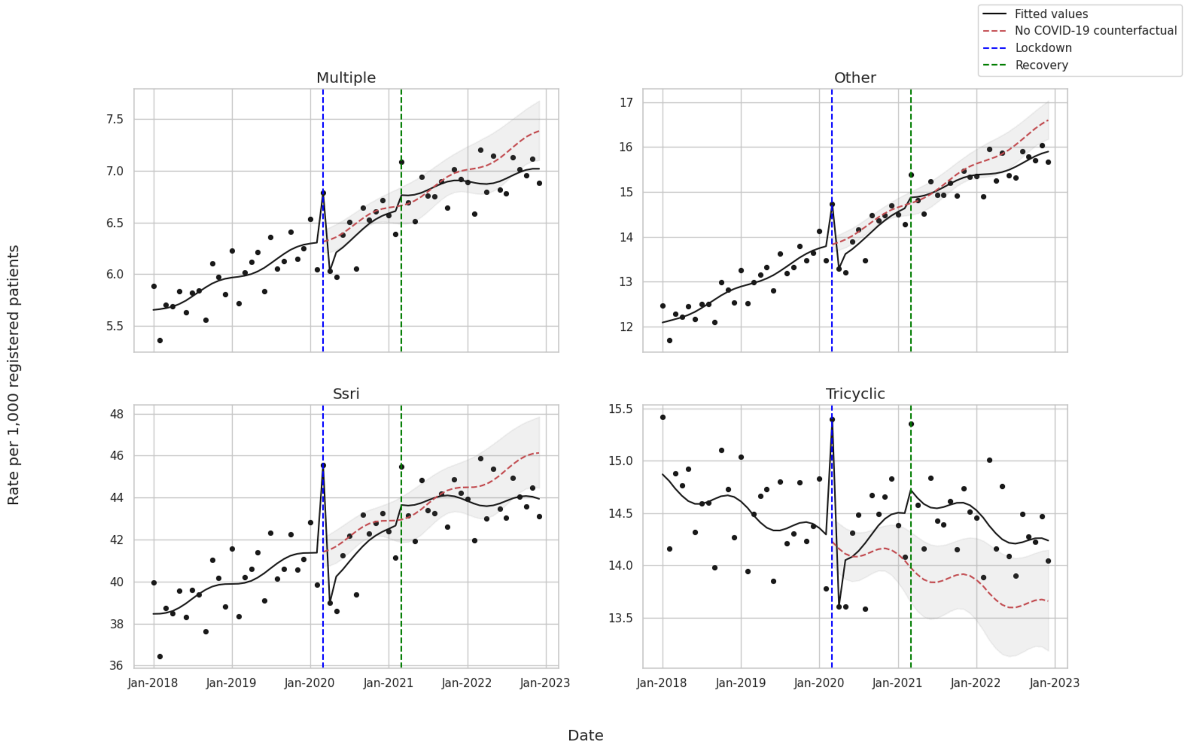

**Figure S7**: (top) Predicted rate of antidepressant prescriptions per 1,000 patients using event level data from OpenPrescribing from September 2018 to December 2022 adjusted for long-term seasonality and trend. The dotted red line is the predicted counterfactual rate, if neither the COVID-19 lockdown or recovery interruptions occurred. (bottom) The relative risk comparing the model fitted rate compared to the no COVID-19 counterfactual from March 2020 though December 2022. A relative risk of 1 represents no change from pre-COVID-19 trends. The overall effect size is computed with the geometric mean of all the time points.

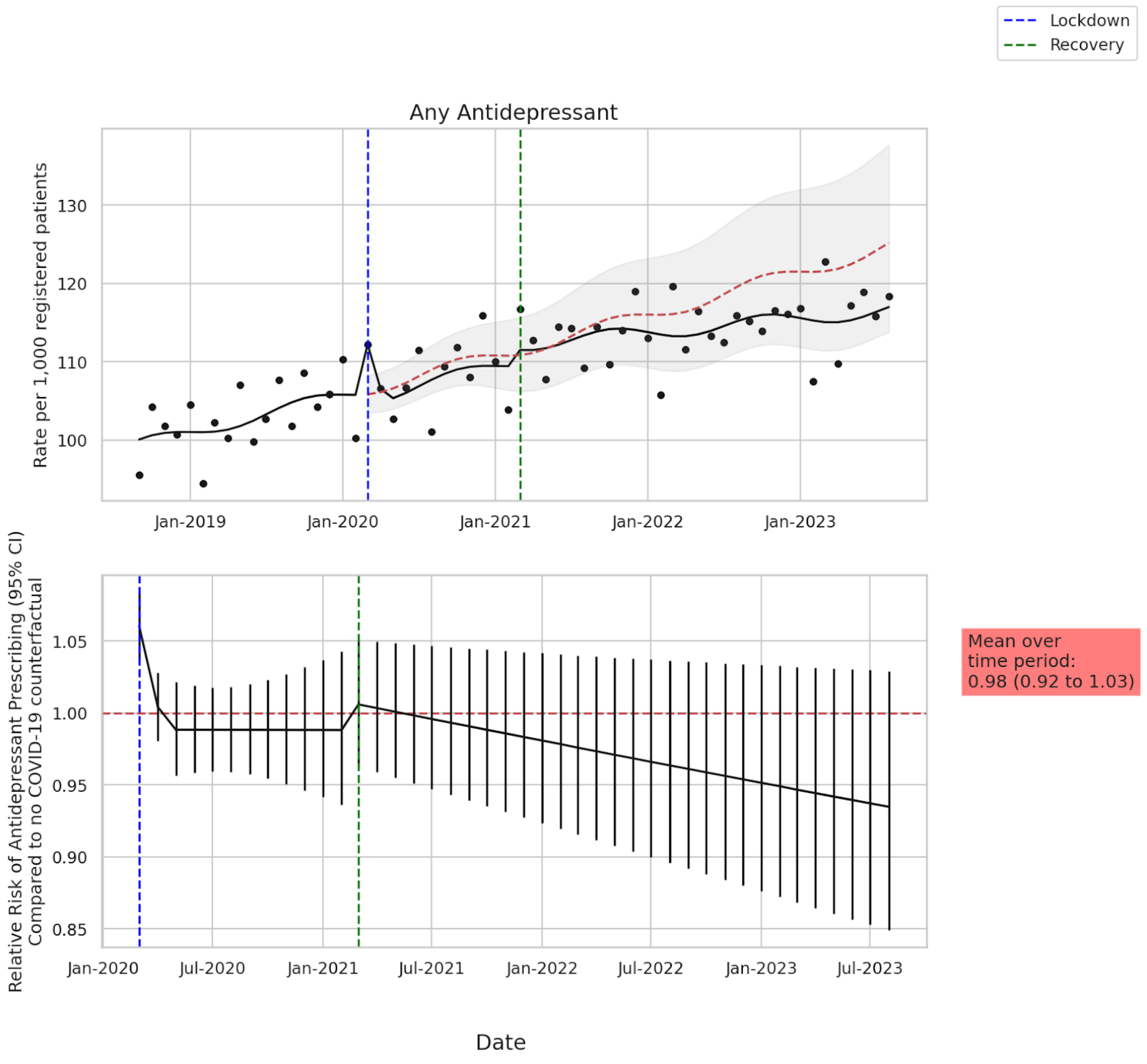

**Figure S8**: Absolute difference in rate between the model predicted rate and the no COVID-19 counterfactual at the start of the Recovery (March 2021) and the end of the study period (December 2022). A rate difference of 0 represents no change from pre-COVID-19 trend.

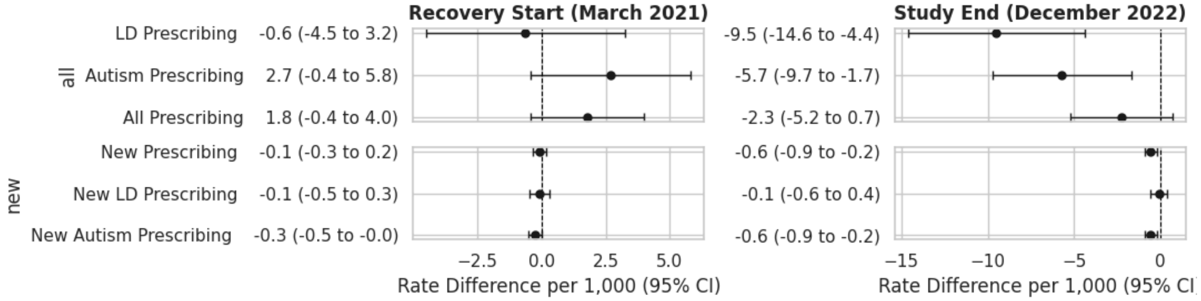

**Figure S9**: Absolute difference in rate between the model predicted rate and the no COVID-19 counterfactual at the end of the study period (December 2022) by demographic subgroup. A rate difference of 0 represents no change from pre-COVID-19 trend.

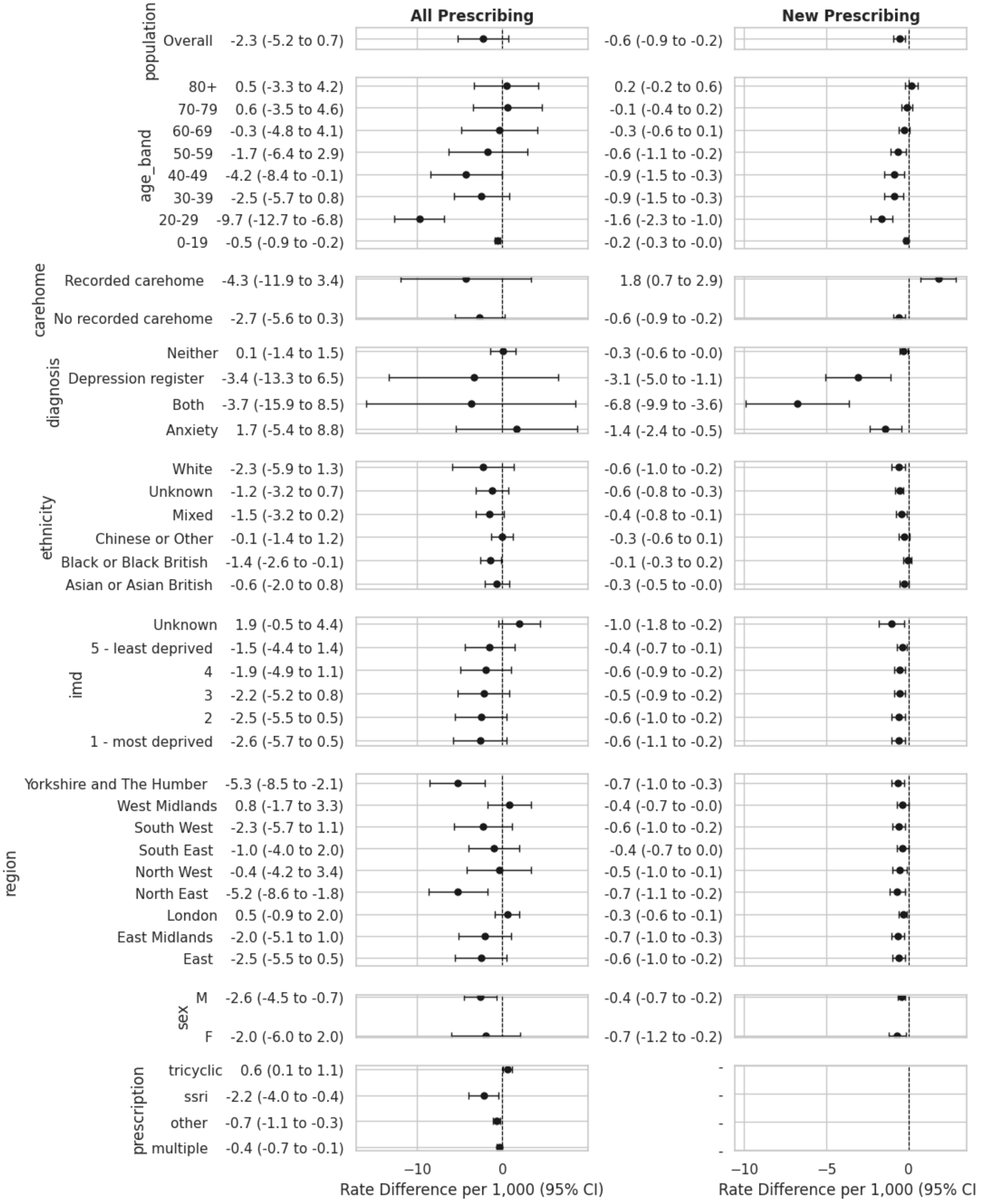
